# Apathy: an underestimated feature in GBA and LRRK2 non-manifesting mutation carriers

**DOI:** 10.1101/2021.07.20.21260819

**Authors:** Ioanna Pachi, Christos Koros, Athina M Simitsi, Dimitra Papadimitriou, Anastasia Bougea, Andreas Prentakis, Nikolaos Papagiannakis, Maria Bozi, Roubina Antonelou, Efthalia Angelopoulou, Ion Beratis, Maria Stamelou, Xenia Geronicola Trapali, Sokratis G. Papageorgiou, Leonidas Stefanis

## Abstract

**Background:** Higher prevalence of motor and non-motor features has been observed in non-manifesting mutation carriers of Parkinson’s Disease (PD) compared to Healthy Controls (HC).

**Objectives:** The aim was to detect the differences between GBA and LRRK2 mutation carriers without PD and HC on neuropsychiatric symptoms.

**Methods:** This is a cross-sectional retrospective study of non-manifesting GBA and LRRK2 mutation carriers and HC enrolled into Parkinson’s Progression Markers Initiative (PPMI). Data extracted from the PPMI database contained: demographics and performance in MoCA scale and MDS-UPDRS scale part 1A (neuropsychiatric symptoms). All six features were treated as both continuous (MDS-UPDRS individual scores) and categorical variables (MDS-UPDRS individual score>0 and MDS-UPDRS individual score=0). Logistic regression analyses were applied to evaluate the association between mutation carrying status and neuropsychiatric symptoms.

**Results:** We found that non-manifesting mutation carriers as a whole (total N=654, GBA: n=285, LRRK2: n=369) were 2.3 times more likely to present apathy compared to HC, even after adjustment for covariates (adjusted OR=2.3, 95% CI=1.1-5.0, p-value=0.027). The effect was mainly driven by GBA mutation carriers (adjusted OR= 2.6, 95% CI=1.1-6.3, p=0.031), while the higher percentage of apathy for LRRK2 carriers compared to HC was marginally non-significant. Other neuropsychiatric symptoms, such as psychotic or depressive manifestations, did not differ between groups.

**Conclusions:** Symptoms of apathy could be present in the premotor period of LRRK2 and, especially, GBA mutation carriers. Longitudinal data, including detailed neuropsychiatric evaluation and neuroimaging, would be essential to further investigate the pathophysiological basis of this finding.

## Introduction

Parkinson’s disease (PD) is the second most common neurodegenerative disorder, clinically characterized by bradykinesia, resting tremor and rigidity, with postural instability ensuing in later stages. The pathological hallmark of PD is the degeneration of dopaminergic substantia nigra pars compacta neurons, although other areas of the brain are also affected [1]. Even though PD is considered a movement disorder, the cardinal motor features are now increasingly recognized as representing the “tip of the iceberg”. The underlying burden of non-motor symptoms encompasses sensory and autonomic dysfunction, such as hyposmia and constipation, sleep disorders, such as REM sleep behavior disorder (RBD), and neuropsychiatric manifestations. Cognitive decline, depression, anxiety, apathy, psychosis and impulsive-compulsive behaviors [ICDs] (including dopamine dysregulation syndrome, DDS) have been well described in early and later stages of PD, even though they tend to be underrecognized in clinical practice [2]. However, they can often precede the motor symptoms by years or decades. In fact, depression and anxiety have been identified several years before the onset of motor features [3-6], while minor hallucinations have been noticed in the premotor era of the illness [7]. Apathy has been reported retrospectively to occur before the onset of motor symptoms [8] and there is limited data about the occurrence of ICDs in the prodromal period.

Since 60-70% of striatal dopaminergic depletion occurs before the onset of motor symptoms, it is of great significance to fully understand the premotor phase of PD, characterize and validate clinical, neuroimaging and genetic biomarkers that would enhance specificity in diagnosis and early intervention. On this basis, a task force of the International Parkinson and Movement Disorder Society (MDS) has developed research criteria for the definition of prodromal PD and an increasing number of studies have focused on “at-risk” populations [3,5,9,10]. Among these populations, non-manifesting gene carriers offer a unique sample for identifying and assessing potential clinical and biological risk characteristics.

Mutations in the LRRK2 and GBA genes have been recognized as the most common genetic alterations associated with increased susceptibility to PD [11-13]. LRRK2-PD is characterized by a typical clinical syndrome of late onset Parkinsonism with a relatively benign course and good response to L-dopa [14]. GBA-PD is characterized by younger age of onset, more severe autonomic dysfunction, more accelerated course, including prominent cognitive decline and more prevalent psychiatric presentations such as depression and anxiety, compared to idiopathic PD [15-17]. There is a growing number of studies examining motor, non-motor and imaging features in non-manifesting carriers of GBA and LRKK2 mutations. In some cohorts a higher prevalence of autonomic dysfunction and subtle motor features in mutation carriers compared to healthy individuals has been observed [18]. However, the presence of neuropsychiatric manifestations during the premotor phase of GBA-PD and LRRK2-PD has not been adequately studied. Several studies have investigated the cognitive profile and the brain activation patterns in asymptomatic carriers and healthy individuals [19,20]. Information is scarce from prospective studies regarding psychiatric manifestations, with comparisons between non-manifesting carriers and healthy individuals.

The primary aim of the current study was to detect the differences in neuropsychiatric symptoms between GBA and LRRK2 mutation carriers without PD and healthy controls. Our hypothesis was that GBA and LRKK2 unaffected carriers would present higher prevalence of psychiatric symptoms, compared to healthy individuals.

## Materials and Methods

### Participants

Data used in the preparation of this study were obtained from the Parkinson’s Progression Markers Initiative (PPMI) database (http://www.ppmiinfo.org/data). PPMI is an ongoing, observational, international and multicenter study designed to identify clinical, imaging and biospecimen biomarkers of PD progression. The protocol, aims and methodology of the study have been published elsewhere [21,22].

In this study, non-manifesting GBA and LRRK2 mutation carriers and healthy controls (HC) were investigated. Inclusion and exclusion criteria of the PPMI study have been presented previously. Briefly, inclusion criteria for non-manifesting mutations carriers (NMMC) were: 1) aged ≥45 years at baseline and 2) a LRRK2 or GBA mutation confirmed by the genetic core. The LRRK2 genetic testing included G2019S and R1441G mutations, while the GBA genetic testing included N370S (in all), and L483P, L444P, IVS2+1, and 84GG (in a subset of participants) mutations. Dual mutation carriers (LRRK2 and GBA) were excluded from this analysis. NMMC were recruited by the participating sites (existing databases) and via a novel strategy that targets first-degree relatives of PD patients of Ashkenazi Jewish ancestry [22]. Participants with a clinical diagnosis of PD based on established diagnostic criteria [0] or with non-available data on demographic, clinical and neuropsychiatric features (MDS-UPDRS Part 1A) were excluded from the current analysis.

All participants provided a written informed consent. The study was approved by the Scientific Board of each PPMI site involved (including the Scientific Board of Eginition Hospital). Data were downloaded on February 5, 2021.

### Clinical and neuropsychiatric evaluation

The clinical assessment battery has been described on the PPMI website and has been published previously. Data extracted from PPMI database included demographic characteristics (age, sex and years of education) and global cognitive testing using the Montreal Cognitive Assessment (MOCA) scale [23]. The neuropsychiatric examination was ascertained by the Movement Disorders Society Unified Parkinson’s Disease Rating Scale (MDS-UPDRS) Part IA that evaluates complex behaviors in six domains: cognitive impairment, hallucinations and psychosis, depressed and anxious mood, apathy and features of Dopamine Dysregulation Syndrome (DDS) [24]. Each item is rated according to the severity by a 5-point scale: 0=normal, 1=slight, 2=mild, 3=moderate and 4=severe. Apart from the total and individual MDS-UPDRS part 1A scores, a categorical variable for each neuropsychiatric feature was computed. Thus, a participant with an MDS-UPDRS part 1A score >0 was grouped as affected, while an individual with a score of 0 was categorized as non-affected.

### Statistical Analysis

Data was analyzed using IBM SPSS version 26.0 (USA). Categorical variables were summarized as absolute numbers and percentages. Continuous variables were presented as mean ± standard deviation (SD).

Primarily, we separated our sample into HC and NMMC. Each neuropsychiatric feature was initially treated as a raw individual MDS-UPDRS score part IA and, secondly, as a categorical variable (individual MDS-UPDRS score>0 and individual MDS-UPDRS score=0). Neuropsychiatric manifestations, MoCA test scores and demographic data including age, sex and years of education were compared between HC and NMMC groups using the Fisher exact test, the χ^2^ test and Student t test as appropriate. Generalized linear models (GLMs) were computed in order to evaluate the association between MDS-UPDRS part IA scores and clinical variables such as age, gender, years of education and mutation carrying status. Additionally, binary logistic regression models were constructed to further examine the association between mutation carrying status (independent variable) and neuropsychiatric symptoms (dependent variables) by adjusting for clinical and demographic characteristics (age, gender, years of education and MoCA score).

Subsequently, we separated the NMMC group into two sub-groups based on their genetic status (GBA or LRRK2), yielding a total of three groups (HC, GBA non-manifesting carriers and LRRK2 non-manifesting carriers). We then performed two exploratory analyses, similar to the main analysis, with genetic status as fixed factor and age, sex, years of education and MOCA scores as clinical confounders.

All statistical tests were two-sided. P < 0.05 is the cut-off point.

## Results

### Demographic and Clinical Characteristics

Of 849 participants, 654 NMMC participants (LRRK2 group: n=369, GBA group: n=285) and 195 HC were included in the analysis. The demographic and clinical characteristics are illustrated in Table 1. There was a predominance of male sex in HC (125 out of 195, 64%), compared to the NMMC group (265 out of 654, 41%) [p<0.001]. NMMC were younger, had higher education level and lower MoCA scores than HC (p<0.001).

**Table 1.**
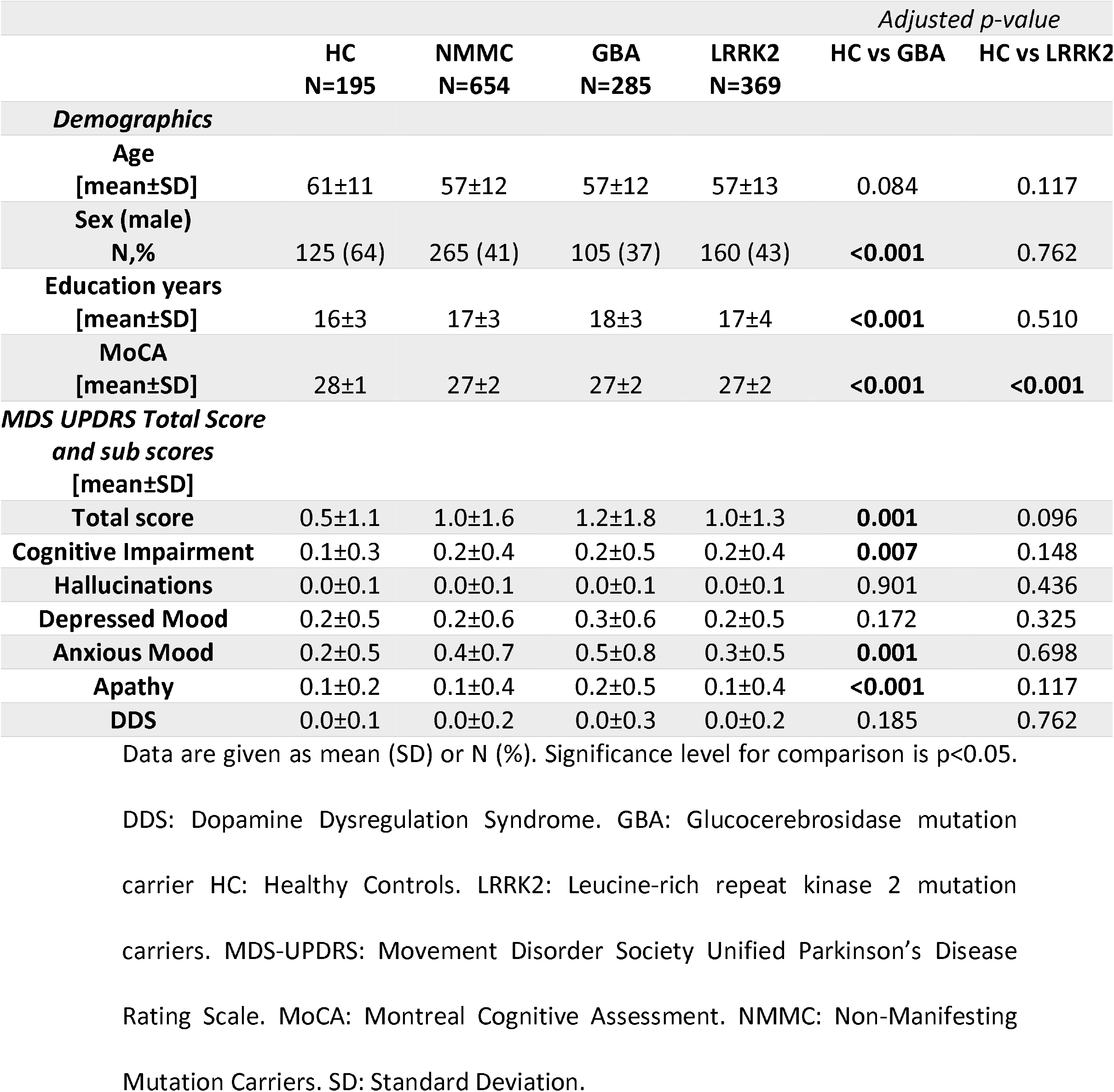
Demographic, Clinical and Neuropsychiatric Features.

### Neuropsychiatric features in NMMC and HC

The majority of participants reported neuropsychiatric features of slight severity, as total and individual MDS-UPDRS Part 1A mean scores were ≤1.0 in both categories (Table 1). NMMC group exhibited higher total MDS-UPDRS part IA scores, compared to HC, after adjusting for clinical covariates (p=0.025) [Table 1].

Regarding individual neuropsychiatric scores, NMMC participants presented higher mean scores in apathy, compared to HC (p=0.027) [Table 2]. The scores of the cognitive and other psychiatric presentations did not differ between the two groups (cognitive impairment: p=0.124, hallucinations: p=0.781, depressive symptoms: p=0.119, anxious symptoms: p=0.249, impulsivity and DDS: p=0.584) [Table 1].

**Table 2.** Neuropsychiatric features.

| N,% | HC<br>N=195 | NMMC<br>N=654 | GBA<br>N=285 | LRRK2<br>N=369 | *Adjusted p-value* |  |
| --- | --- | --- | --- | --- | --- | --- |
|  |  |  |  |  | HC vs GBA | HC vs LRRK2 |
| **Cognitive Impairment** | 19 (10) | 111 (17) | 52 (18) | 59 (16) | 0.095 | 0.159 |
| **Hallucinations** | 1 (1) | 4 (1) | 1 (0.4) | 3 (1) | 0.372 | 0.453 |
| **Depressed Mood** | 24 (12) | 114 (17) | 54 (19) | 60 (16) | 0.209 | 0.390 |
| **Anxious Mood** | 38 (20) | 191 (29) | 99 (35) | 92 (25) | 0.167 | 0.561 |
| **Apathy** | 9 (5) | 62 (10) | 31 (11) | 31 (8) | **0.031** | 0.075 |
| **DDS** | 3 (2) | 12 (2) | 6 (2) | 6 (2) | 0.462 | 0.951 |

Data are given as N (%). Generalized Linear Models adjusted for age, sex and years of education. Significance level for comparison is $p < 0.05$ . DDS: Dopamine Dysregulation Syndrome. GBA: Glucocerebrosidase mutation carrier. HC: Healthy Controls. LRRK2: Leucine-rich repeat kinase 2 mutation carriers. NMMC: Non-Manifesting Mutation Carriers.

When treated as categorical variable, apathy was more prevalent in NMMC group (62 out of 654, 10%), compared to HC (9 out of 185, 5%), using GLMs (p=0.026) [Table 2]. Binary regression analyses revealed a double probability of presenting symptoms of apathy in case of mutation carrying status compared to HC (adjusted OR=2.3, 95% CI=1.1-5.0, p=0.027) [Table 3]. No significant difference was detected between the two categories in other neuropsychiatric features [Table 3].

**Table 3.** Neuropsychiatric Features in NMMC and HC.

| **Neuropsychiatric Features**<br>**N,%** | **HC**<br>**N=195** | **NMMC**<br>**N=654** | **Unadjusted OR**<br>**[95% CI, p-value]** | **Adjusted OR**<br>**[95% CI, p-value]** |
| --- | --- | --- | --- | --- |
| **Cognitive Impairment** | 19 (10) | 111 (17) | 1.9 [1.1-3.2, 0.015] | 1.7 [1.0-2.9, 0.066] |
| **Hallucinations** | 1 (1) | 4 (1) | 1.2 [0.1-10.7, 0.874] | 1.2 [0.1-14.2, 0.896] |
| **Depressed Mood** | 24 (12) | 114 (17) | 1.5 [0.9-2.4, 0.090] | 1.4 [0.8-2.3, 0.203] |
| **Anxious Mood** | 38 (20) | 191 (29) | 1.7 [1.2-2.5, 0.008] | 1.3 [0.8-1.9, 0.292] |
| **Apathy** | **9 (5)** | **62 (10)** | **2.2 [1.1-4.4, 0.035]** | **2.3 [1.1-5.0, 0.027]** |
| **DDS** | 3 (2) | 12 (2) | 1.2 [0.3-4.3, 0.783] | 1.0 [0.2-4.0, 0.992] |

Data are given as N (%). Logistic regression analyses adjusted for age, sex and years of education. Significance level for comparison is $p < 0.05$ . CI: Confidence Interval. DDS: Dopamine Dysregulation Syndrome. HC: Healthy Controls. NMMC: Non-Manifesting Mutation Carriers. OR: Odds Ratio.

### GBA non-manifesting carriers VS HC

Higher prevalence of female sex, higher education level and poorer MOCA scores were detected in the GBA group (p<0.001) [Table 1].

Total MDS-UPDRS Part 1A score and individual scores of cognitive decline, anxiety and apathy were found to be higher in the GBA group than in HC (p=0.001, p=0.007, p=0.001, p<0.001 respectively).

After performing binary regression analyses, there was an approximate 3-fold risk of developing features of apathy in GBA group, compared to HC (adjusted OR= 2.6, 95% CI=1.1-6.3, p=0.031). No significant difference was detected between the two groups in other neuropsychiatric symptoms, after adjusting for clinical and demographic factors (Table 4).

**Table 4.** Neuropsychiatric Features in GBA non-manifesting carriers and HC.

| **Neuropsychiatric Features**<br>**N,%** | **HC**<br>**N=195** | **GBA**<br>**N=285** | **Unadjusted OR**<br>**[95% CI, p-value]** | **Adjusted OR**<br>**[95% CI, p-value]** |
| --- | --- | --- | --- | --- |
| **Cognitive Impairment** | 19 (10) | 52 (18) | 2.1 [1.2-3.6, 0.011] | 1.7 [0.9-3.4, 0.095] |
| **Hallucinations** | 1 (1) | 1 (0.4) | 0.7 [0.0-11.0, 0.788] | 0.1 [0.0-31.3, 0.372] |
| **Depressed Mood** | 24 (12) | 54 (19) | 1.7 [1.0-2.8, 0.054] | 1.5 [0.8-2.8, 0.209] |
| **Anxious Mood** | 38 (20) | 99 (35) | 2.2 [1.4-3.4, <0.001] | 1.4 [0.9-2.4, 0.167] |
| **Apathy** | 9 (5) | 31 (11) | **2.5 [1.2-5.4, 0.018]** | **2.6 [1.1-6.3, 0.031]** |
| **DDS** | 3 (2) | 6 (2) | 1.4 [0.3-5.6, 0.654] | 2.0 [0.3-12.3, 0.462] |

Data are given as N (%). Logistic regression analyses adjusted for age, sex and years of education. Significance level for comparison is $p < 0.05$ . CI: Confidence Interval. DDS: Dopamine Dysregulation Syndrome. GBA: Glucocerebrosidase mutation carrier HC: Healthy Controls. OR: Odds Ratio.

### LRRK2 non-manifesting carriers VS HC

MoCA scores were significantly lower in LRRK2 carriers, compared to HC (p<0.001). The six neuropsychiatric presentations, either treated as continuous or categorical variables, did not differ between the two groups (Tables 1, 2, 5).

**Table 5.**
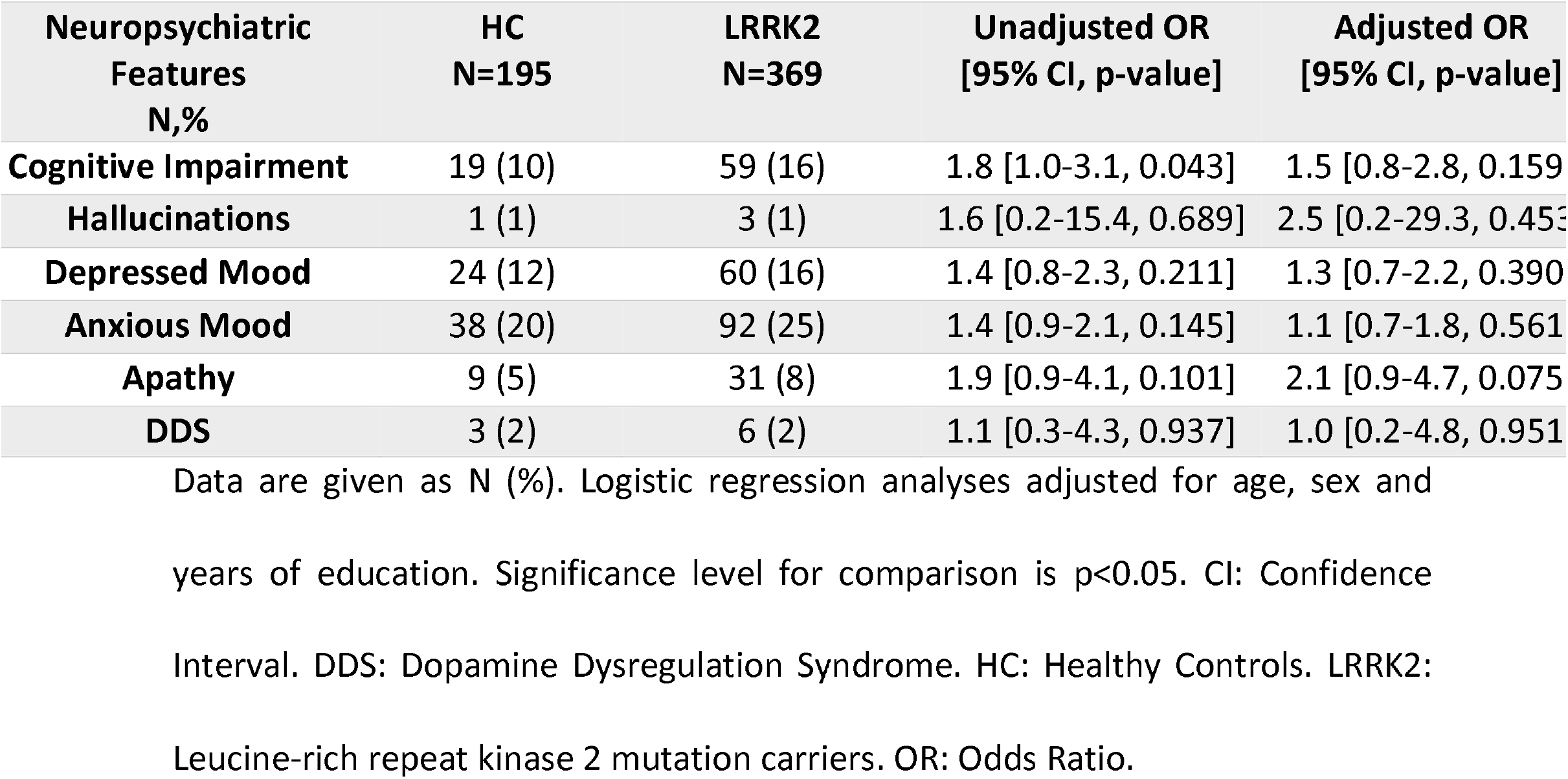
Neuropsychiatric Features in LRRK2 non-manifesting carriers and HC.

| **Neuropsychiatric Features**<br>**N,%** | **HC**<br>**N=195** | **LRRK2**<br>**N=369** | **Unadjusted OR**<br>**[95% CI, p-value]** | **Adjusted OR**<br>**[95% CI, p-value]** |
| --- | --- | --- | --- | --- |
| **Cognitive Impairment** | 19 (10) | 59 (16) | 1.8 [1.0-3.1, 0.043] | 1.5 [0.8-2.8, 0.159] |
| **Hallucinations** | 1 (1) | 3 (1) | 1.6 [0.2-15.4, 0.689] | 2.5 [0.2-29.3, 0.453] |
| **Depressed Mood** | 24 (12) | 60 (16) | 1.4 [0.8-2.3, 0.211] | 1.3 [0.7-2.2, 0.390] |
| **Anxious Mood** | 38 (20) | 92 (25) | 1.4 [0.9-2.1, 0.145] | 1.1 [0.7-1.8, 0.561] |
| **Apathy** | 9 (5) | 31 (8) | 1.9 [0.9-4.1, 0.101] | 2.1 [0.9-4.7, 0.075] |
| **DDS** | 3 (2) | 6 (2) | 1.1 [0.3-4.3, 0.937] | 1.0 [0.2-4.8, 0.951] |

Data are given as N (%). Logistic regression analyses adjusted for age, sex and years of education. Significance level for comparison is $p < 0.05$ . CI: Confidence Interval. DDS: Dopamine Dysregulation Syndrome. HC: Healthy Controls. LRRK2: Leucine-rich repeat kinase 2 mutation carriers. OR: Odds Ratio.

## Discussion

In this study we found that among six neuropsychiatric features, apathy was significantly more prevalent in the combined group of GBA and LRRK2 mutation carriers, compared to HC. In fact, mutation carrying status was associated with a more than double probability of manifesting apathy, compared to controls. This difference was mainly driven by the GBA group, as GBA non-manifesting carriers were almost three times more likely to present apathy compared to HC. LRRK2 asymptomatic carriers and healthy individuals presented similar performance in neuropsychiatric evaluation of the MDS-UPDRS scale, although there was a tendency for apathy again to be more prevalent in the LRRK2 carrier group. Hallucinations, depression, anxiety and DDS did not differ between NMMC and HC.

To the best of our knowledge, this is the first study comparing the prevalence of apathy between GBA & LRRK2 non-manifesting carriers and healthy individuals. In a large longitudinal study of GBA mutation-positive individuals without PD that evaluated the progression of prodromal features over a two-year period, apathy was not investigated as a potential clinical marker during premotor phase [25]. In recent studies that focused on cognitive, behavioural and psychiatric disturbances including depression and anxiety in non-manifesting GBA and / or LRRK2 mutation carriers, apathy was not individually examined [18,20,26-31].

On the other hand, cohorts of prodromal PD have addressed the issue of behavioural disturbances including apathy in the premotor PD period. Fereshtehnejad et al. (2019) have traced the evolution of early motor and non-motor features of synucleinopathy from the stage of idiopathic RBD until defined PD or dementia with Lewy bodies (DLB) [8]. They observed that apathy preceded PD or DLB diagnosis by six years. Moreover, apathy, as a distinct entity from depression and cognitive impairment, has been present in approximately 50% of an idiopathic RBD population, albeit relatively underrecognized [32]. A potential mechanism for the high prevalence of apathy in RBD samples has been provided by Barber et al (2018), who suggested that this is due to serotoninergic dysfunction in the dorsal raphe nucleus, which is in close anatomical proximity to the locus coeruleus/subcoeruleus complex, an important player in the regulation of REM sleep [33]. Interestingly, a strong association between GBA mutations and RBD has been reported [25,34-36]. Therefore, there might be a common network dysfunction linking apathy and RBD in GBA carriers. Since the role of dopaminergic, serotoninergic or other neurotransmitter system dysfunction has not yet been clarified in asymptomatic GBA carriers, further studies of GBA prodromal cohorts are required to ascertain the pathophysiological basis responsible for apathy in this setting.

The pathophysiological basis of apathy is still unknown. Regarding PD, results of structural and functional studies have failed to provide a unique anatomical pattern underlying apathy, even though it has been associated with deficits of prefrontal-basal ganglia circuits [37,38]. Richter et al (2018) first have shown that, apart from depressive features, symptoms of apathy in sporadic PD patients are correlated to brainstem raphe signal alterations in transcranial sonography [39]. These changes in the micro-architecture of the raphe nucleus may reflect serotoninergic deficiency, which could play a role in the pathophysiology of apathy. Concerning especially GBA-PD, morphologic alterations of the midbrain raphe have been more prevalent, compared to idiopathic PD and they may correspond to more frequent and severe manifestation of neuropsychiatric disturbances [17]. However, since the design of these studies was cross-sectional, the temporal association between the reduced signal in raphe nuclei and apathy was not investigated. In Braak stage 2 and 3, pontine regions are affected by accumulation of Lewy body pathology, before neurons in the substantia nigra are implicated [40]. Therefore, it could be hypothesized that alpha synuclein pathology in serotoninergic areas of raphe nuclei could produce subtle symptoms of apathy experienced in the prodromal phase of PD, including GBA non-manifesting carriers, as shown in the current study.

Few data are available on LRRK2 prodromal studies [20,29,30,31,41]. Non-motor symptoms are relatively mild in LRKK2 mutation carriers without PD. Compared to non-carriers, higher scores in motor scales and reduced nigrostriatal dopamine innervation have been observed in LRRK2 asymptomatic carriers. Wile et al (2017) first reported an increased serotonin transporter binding in the striatum, brainstem and hypothalamus of LRRK2 mutation carriers without PD [42]. Unfortunately, in this study, the correlation of elevated serotonin binding to neuropsychiatric disturbances was not part of the analysis. In our study, non-manifesting LRRK2 carriers did not present any statistical difference in apathy, compared to controls, although there was a trend favoring the LRRK2 carrier group. Taken together, these findings may raise the question: could elevated serotonin binding in brainstem regions of LRRK2 non-manifesting carriers be associated with the lower prevalence of apathetic symptoms compared to other forms of prodromal PD? In the case that future findings support this idea, then the theory that serotoninergic deficits may facilitate presentations of apathy in the prodromal phase of PD would be further strengthened. Larger cohorts with longitudinal observations combining both imaging data and monitoring of neuropsychiatric disturbances will be needed to assess this hypothesis in non-manifesting GBA and LRRK2 carriers.

In our study, depression, hallucinations, anxiety and ICDs did not differ between NMMC and HC. Our findings are in line with recent literature, even though we used a single part of the MDS-UPDRS scale instead of sensitive and detailed scales. A recent study of GBA and LRRK2 non-manifesting carriers recruited in PPMI reported no difference in Geriatric Depression Scale (GDS) scores, Questionnaire for Impulsive-Compulsive Disorders in PD (QUIP) scores and State trait anxiety scores, compared to non-carriers [18]. Furthermore, similarly to that study, we noticed that total MDS-UPDRS score part 1A was significantly higher in both asymptomatic mutation carriers compared to healthy individuals. Mirelman et al. (2018) implemented the MDS criteria for prodromal PD in healthy G2019S-LRRK2 carriers [30]. The comparison between LRRK2 carriers and healthy non-carriers revealed a non-significant trend for increased depression (p=0.071) in the LRRK2 group. In a retrospective analysis of GBA-carriers, LRKK2-carriers and idiopathic PD, features of anxiety and depression were indistinguishable between the groups [28]. Taken together, our outcomes are in line with recent literature findings on the premotor period of LRRK2 mutation carriers. Data on psychotic features in prodromal period of GBA and LRRK2 mutation carriers has been very limited. In our study, slight features of perceptual abnormalities were noticed in up to 1% of both healthy individuals and asymptomatic mutation carriers. This could mean that psychotic symptoms are either truly rare phenomena in the prodromal phase of PD or that more sensitive evaluation scales are needed to detect them. Lastly, poorer performance in MoCA scores was detected in mutation carriers compared to HC, despite the fact that the individual item of cognitive impairment in MDS-UPDRS scale did not differ between the two groups. This finding is in accordance with previously published results in cohorts of non-manifesting carriers of LRRK2 and GBA mutations [18,20].

Our results, though, may have been influenced by some weaknesses. The main limitation of this study is the use of a single item from the MDS-UPRDS scale, part 1A, to document the presence of each neuropsychiatric feature. This type of questionnaire was mainly applied for screening reasons, so it may have failed to detect mild neuropsychiatric features including depression, hallucinations, anxiety, apathy and ICDs-DDS. Therefore, these features might have been underestimated. An exception to this was cognitive impairment, as MoCA scale was additionally implemented. Secondly, the cross-sectional design of the current study provided only baseline observations. Comprehensive longitudinal follow-up neuropsychiatric examinations would be crucial to confirm the findings of this study and investigate the potential underlying mechanisms. Finally, higher MDS-UPDRS scores in both asymptomatic mutation carriers could be driven by the bias of examiners who, in most cases, were aware of the participant’s genetic status. In that case, outcomes and correlations found in this study could be affected as well. However, the consistency of our results with recent literature suggests that a pathological and biological basis may exist rather than an ascertainment bias. Future investigations that also include data on neuroimaging biomarkers could help clarify the pathophysiological basis of our outcomes.

On the other hand, our study has several strengths. The novelty of our study is the identification of mild symptoms of apathy in the premotor phase of non-manifesting carriers, especially in case of the GBA group. We took under consideration potential confounders such as age and sex that were associated with both mutation carrying status and neuropsychiatric features. Moreover, the size of our cohort was substantially large and representative of GBA and LRRK2 non-manifesting carriers. These data have provided the opportunity to evaluate in parallel clinical characteristics of asymptomatic mutation carriers and healthy controls, with the same scope of assessments, as they were all part of the same protocol and underwent the same clinical examination.

To summarize, our results have shown increased prevalence of apathy collectively in GBA and LRRK2 non-manifesting carriers, compared to non-carriers. Interestingly, GBA carriers have mainly contributed to this finding, due to their nearly three-fold risk of presenting features of apathy compared to healthy individuals. The implementation of sensitive neuropsychiatric scales in longitudinal cohorts of asymptomatic GBA and LRRK2 mutations carriers would be essential to confirm these outcomes and better characterize the presence of psychiatric features in the prodromal period of PD.

## Data Availability

Data used in the preparation of this study were obtained from the Parkinson s Progression Markers Initiative (PPMI) database over the period 2014-2019 (http://www.ppmiinfo.org/data).

## Acknowledgment

We are grateful to the patients and relatives for their participation in this project. We thank volunteer healthy controls who contributed samples for this study. This work was supported by the Michael J. Fox Foundation for Parkinson’s Research (https://www.ppmi-info.org/data).

## Authors’ Role

1. Research project: A. Conception, B. Organization, C. Execution.
2. Statistical Analysis: A. Design, B. Execution, C. Review and Critique.
3. Manuscript: A. Writing of the first draft, B. Review and Critique.

| Name | Role |
| --- | --- |
| Ioanna Pachi | 2A,2B,3A |
| Christos Koros | 2C, 3B |
| Athina M Simitsi | 2C, 3B |
| Dimitra Papadimitriou | 2C, 3B |
| Anastasia Bougea | 2C, 3B |
| Andreas Prentakis | 2C, 3B |
| Nikolaos Papagiannakis | 2C, 3B |
| Maria Bozi | 2C, 3B |
| Roubina Antonellou | 2C, 3B |
| Efthalia Angelopoulou | 2C, 3B |
| Ion Beratis | 2C, 3B |
| Maria Stamelou | 2C, 3B |
| Xenia Geronicola Trapali | 2C, 3B |
| Sokratis G Papageorgiou | 2C, 3B |
| Leonidas Stefanis | 1A, 1B,3B |

## Financial Disclosures of all authors

C. Koros received funding from the Michael J. Fox Foundation for his participation in Parkinson’s Progression Markers Initiative (PPMI). A.M/ Simitsi received funding from the Michael J Fox Foundation for her participation in PPMI. I. Beratis received funding from the Michael J Fox Foundation for his participation in PPMI. M. Stamelou serves on the editorial boards of Movement Disorders Journal and Frontiers in Movement Disorders and receives research support from the Michael J. Fox Foundation (PPMI). L. Stefanis has received the following grants: MULTISYN European Program (EU, FP7-HEALTH.2013.1.2-1, number 602646), PPMI (supported by the Michael J. Fox Foundation), IMPRIND-IMI2 Number 116060 (EU, H2020), “PBMC and urine collection in LRRK2 and idiopathic PD” Grant by the Michael J. Fox Foundation (collaborator). I. Pachi, D. Papadimitriou, A. Bougea, A. Prentakis, N. Papagiannakis, M. Bozi, R. Antonellou, E. Angelopoulou, X. G. Trapali and S. G. Papageorgiou report no disclosures relevant to the manuscript.

## Notes

### Competing Interest Statement

C.K. received funding from the Michael J. Fox Foundation for his participation in Parkinson s Progression Markers Initiative (PPMI). A.S. received funding from the Michael J Fox Foundation for her participation in PPMI. I.B. received funding from the Michael J Fox Foundation for his participation in PPMI. M.S. serves on the editorial boards of Movement Disorders Journal and Frontiers in Movement Disorders and receives research support from the Michael J. Fox Foundation (PPMI). L.S. has received the following grants: MULTISYN European Program (EU, FP7‐HEALTH.2013.1.2‐1, number 602646), PPMI (supported by the Michael J. Fox Foundation), IMPRIND‐IMI2 Number 116060 (EU, H2020), PBMC and urine collection in LRRK2 and idiopathic PD Grant by the Michael J. Fox Foundation (collaborator). I.P, D.P., A.B., A.P., N.P., M.B., R.A., E.A., X.G.T. and S.G.P report no disclosures relevant to the manuscript.

### Funding Statement

: C.K. received funding from the Michael J. Fox Foundation for his participation in Parkinson s Progression Markers Initiative (PPMI). A.S. received funding from the Michael J Fox Foundation for her participation in PPMI. I.B. received funding from the Michael J Fox Foundation for his participation in PPMI. M.S. serves on the editorial boards of Movement Disorders Journal and Frontiers in Movement Disorders and receives research support from the Michael J. Fox Foundation (PPMI). L.S. has received the following grants: MULTISYN European Program (EU, FP7‐HEALTH.2013.1.2‐1, number 602646), PPMI (supported by the Michael J. Fox Foundation), IMPRIND‐IMI2 Number 116060 (EU, H2020), PBMC and urine collection in LRRK2 and idiopathic PD Grant by the Michael J. Fox Foundation (collaborator). I.P, D.P., A.B., A.P., N.P., M.B., R.A., E.A., X.G.T. and S.G.P report no disclosures relevant to the manuscript.

